# COVID-19 Infection Is Associated with Poor Outcomes in Patients with Intracerebral Hemorrhage

**DOI:** 10.1101/2023.05.03.23289488

**Authors:** Daniela Renedo, Audrey C. Leasure, Rebecca Young, Cyprien Rivier, Brooke Alhanti, Brian Mac Grory, Steven R. Messe, Matthew Reeves, Ameer E. Hassan, Lee Schwamm, Adam De Havenon, Charles C. Matouk, Kevin N. Sheth, Guido J. Falcone

**Author notes:** **Corresponding author** Guido J. Falcone, MD ScD MPH, 100 York Street, Office 111, New Haven, CT 06511, USA. Co-first authors (Daniela Renedo, Audrey C. Leasure). Jointly supervised this work (Kevin N. Sheth, Guido J. Falcone).

## Abstract

**Background:** Patients with ischemic stroke and concomitant coronavirus 2019 (COVID-19) infection have worse outcomes than those without this infection. However, research on the impact of COVID-19 infection on outcomes following hemorrhagic stroke remains limited. We aim to study whether concomitant COVID-19 infection leads to worse outcomes in spontaneous intracerebral hemorrhage (ICH).

**Design:** We conducted an observational study using data from Get With The Guidelines® Stroke, an ongoing, multi-center, nationwide quality assurance registry.

**Methods:** We implemented a two-stage design: first, we compared outcomes of ICH patients with and without COVID-19 infection admitted during the pandemic (from March 2020 to February 2021). Second, we compared the same outcomes between ICH patients admitted before (March 2019 to February 2020) and during (March 2020 and February 2021) the pandemic. Main outcomes were poor functional outcome (defined as a modified Rankin Scale of 4 to 6 [mRS] at discharge), mortality and discharge to skilled nursing facility (SNF) or hospice.

**Results:** The first stage included 60,091 COVID-19-negative and 1,326 COVID-19-positive ICH patients. In multivariable analyses, ICH patients with versus without COVID-19 infection had 68% higher odds of poor outcome (OR 1.68, 95%CI 1.41-2.01), 51% higher odds of mortality (OR 1.51, CI 1.33-1.71) and 66% higher odds of being discharged to a SNF/hospice (OR 1.66, 95%CI 1.43-1.93). The second stage included 62,743 pre-pandemic and 64,681 intra-pandemic ICH cases. In multivariable analyses, ICH patients admitted during versus before the COVID-19 pandemic had a 10% higher odds of poor outcome (OR 1.10, 95%CI 1.07-1.14), 5% higher mortality (OR 1.05, 95%CI 1.02-1.08) and no significant difference in the risk of being discharged to SNF/hospice (OR 0.93, 95%CI 0.90-0.95).

**Conclusions:** The pathophysiology of the COVID-19 infection and changes in healthcare delivery during the pandemic played a role in worsening outcomes in this patient population. Further research is needed to identify these factors and understand their effect on the long-term outcome.

## INTRODUCTION

COVID-19 can significantly worsen vascular diseases through activation of the coagulation cascade, activation of platelet-related pathways, and exacerbation of inflammatory responses.^1–3^ Several studies have shown that COVID-19 leads to higher risk of, and worse outcomes after, ischemic stroke. ^4–7^ However, the role of COVID-19 in hemorrhagic stroke remains understudied, largely due to the difficulty of attaining the necessary sample size to meaningfully study this question in a stroke subtype that is relatively rare. A previous study showed that patients with spontaneous ICH or SAH and comorbid COVID infection were more likely have diabetes been obese and to have higher rates of death when compared with controls.^8^

To overcome these limitations, we leveraged the American Heart Association Get With The Guidelines® (GWTG)-Stroke, an ongoing national registry of patients hospitalized for stroke.^9^ Over 2,000 hospitals across the country contribute data to GWTG, significantly facilitating new discoveries in areas of stroke research were the stroke type, exposure or outcome are rare or infrequent.^10,11,^ Leveraging this unique platform, we conducted an observational study aimed at characterizing the clinical evolution of patients with primary, non-traumatic intracerebral hemorrhage (ICH) during the COVID-19 pandemic.

## METHODS

### Study design

We performed a retrospective, observational, cohort study using data from ICH patients enrolled in GWTG-Stroke, an ongoing registry that currently includes over 2,000 hospitals and over than 4 million patients.^9–12^ Each participating hospital received either human research approval to enroll cases without individual patient consent under the common rule, or a waiver of authorization and exemption from subsequent review by their institutional review board (IRB). In addition, the institution-wide IRB for the American Heart Association determined that this study is exempt from IRB oversight. Deidentified patient data from participant hospitals is entered into the GWTG-Stroke database. Variables collected include demographic characteristics, medical history, acute treatment (including intravenous thrombolysis and endovascular thrombectomy), clinical outcomes, mortality and discharge destination. Using these data, we conducted a 2-stage study: first, we compared functional outcomes and mortality of ICH patients with and without COVID-19 infection admitted during the pandemic (from March 2020 to February 2021); second, we compared the same outcomes between pre-pandemic ICH patients (admitted between March 2019 and February 2020) and ICH patients admitted during the pandemic (admitted between March 2020 and February 2021).

### Ascertainment of ICH cases

In GWTG–Stroke, trained hospital personnel ascertained ICH cases of patients 18 years of age or older using a combination of clinical data available in the patients’ medical chart, review of neuroimaging and review of discharge International Classification of Diseases (ICD) 9 and 10 codes. Data is entered in the Patient Management Tool, a web-based tool designed to facilitate and standardize the collection of relevant data for each case.^10^

### Ascertainment of COVID-19

Patients were considered COVID-19 positive if they were positive at the time of admission or at any point during the hospitalization. A dedicated variable for COVID-19 status was added to the GWTG–Stroke platform on April 1st, 2020.^13^

### Outcomes

Our primary outcome of interest was post-ICH functional status, evaluated through the modified Rankin scale (mRS), a 6-category scale where 0 = complete recovery with no residual symptoms; 1 = no significant disability, able to carry out all usual activities despite some symptoms; 2 = slight disability, able to look after own affairs without assistance, but unable to carry out all previous activities; 3 = moderate disability, requires some help but able to walk unassisted; 4 = moderately severe disability, unable to attend to own bodily needs without assistance and unable to walk unassisted; 5 = severe disability, requires constant nursing care and attention and is bedridden/incontinent; and 6 = death. Secondary outcomes included in-hospital death, and discharge disposition (dichotomized as skilled nursing facility or hospice versus home, inpatient rehabilitation facility, intermediate care or long-term care).

### Covariates

Other variables utilized in this study included demographic characteristics (age, sex and race/ethnicity), medical history (prior coronary artery disease, atrial fibrillation, heart failure, stroke and chronic kidney failure), vascular risk factors (hypertension, diabetes, hypercholesterolemia, smoking, sleep apnea and alcohol/drug abuse), pre-ICH medications (antiplatelets and anticoagulants), pre-ICH functional status; baseline physiological variables (blood pressure, heart rate and respiratory rate) and information related to the admission (arrival via emergency medical services, arrival on or off-hours, admission NIHSS score, time from symptom onset to intravenous tPA administration and endovascular thrombectomy, baseline laboratory values).

### Statistical Analysis

We used counts (percentages [%]) to describe discrete variables and mean (standard deviation [SD]) to describe continuous variables. For unadjusted associations, we used chi-square or Wilcoxon rank-sum tests, as appropriate. We used multivariable logistic regression models when evaluating post-ICH functional status, in hospital mortality, and discharge disposition. Multivariable models were adjusted for universal confounders (patient age, sex and race/ethnicity), vascular risk factors (hypertension, hyperlipidemia, diabetes, smoking and obesity), comorbidities (prior coronary artery disease, atrial fibrillation, heart failure, stroke and chronic kidney failure), relevant medications (antiplatelets and anticoagulants) and pre-ICH functional status. In a sensitivity analysis, we restricted the cohort to ICH patients who were only admitted to comprehensive stroke centers. All analysis used a complete case approach, where study participants with available data for the outcome, exposure and covariates of interest are included in the analysis. Statistical analyses were conducted using SAS 9.4.

We adhered to the STROBE (Strengthening the Reporting of Observational Studies in Epidemiology) to ensure transparent and complete reporting of our study. To this end, we used the checklists to guide the reporting of our study design, methods, results, and conclusions.^14^

## Results

### Differences between ICH patients with and without COVID-19 infection admitted during the pandemic

The first stage of this study, focused on comparing ICH patients with and without concomitant COVID-19 admitted during the pandemic, evaluated a total of 61,417 ICH patients (mean age 67.4 [SD 15.3], female sex n= 28,923 [46.1%]), including 1,326 with concomitant infection and 60,091 without (Table 1). Compared to ICH patients who were not infected, those with concomitant infection were younger (65 versus 69 years of age) and more likely to be black (21 versus 18%) or Hispanic (18 versus 10%, all p<0.05; Table 1).

**Table 1.** Description and comparison of patient’s baseline characteristics among ICH patients who are COVID-19-positive compared to those with no documented COVID-19 infection.

| Variable | Overall | Positive for COVID-19 | Negative for COVID-19 | P-value |
| --- | --- | --- | --- | --- |
|  | **61,417** | **N=1,326** | **N=60,091** |  |
| **Patient Demographics, n (%)** |  |  |  |  |
| Age, mean (SD) | 67.4 (15.3) | 64.1(15.9) | 67.4(15.2) | <.0001 |
| Female, | 28,923 (46.1%) | 589 (44.4%) | 28,334 (46.1%) | 0.2 |
| Race/Ethnicity |  |  |  | <.0001 |
| White | 37,603(59.9%) | 625(47.1%) | 36,978(60.2%) |  |
| Black | 11,234(17.9%) | 281(21.1%) | 10,953(17.8%) |  |
| Hispanic | 64,42(10.2%) | 244(18.4%) | 6,198(10.1%) |  |
| Asian | 33,94(5.4%) | 64(4.8%) | 3,330(5.4%) |  |
| Other | 40,44(6.4%) | 112(8.4%) | 3,932(6.4) |  |
| Missing | 26 (0.04%) | 0 (0.0%) | 26 (0.04%) |  |
| **Vascular risk factors** |  |  |  |  |
| Hypertension | 45,683 (73.4%) | 970 (73.2%) | 44,713 (73.4%) | 0.901 |
| Dyslipidemia | 24,109 (38.7%) | 488 (36.8%) | 23,621 (38.7%) | 0.154 |
| Diabetes Mellitus | 16,515 (26.5%) | 451 (34.0%) | 16,064 (26.3%) | <.0001 |
| Obesity/Overweight | 18,809 (30.2%) | 415 (31.3%) | 18,394 (30.2%) | 0.370 |
| Smoker | 8,277 (13.3%) | 141 (10.6%) | 8,136 (13.3%) | 0.004 |
| **Comorbidities** |  |  |  |  |
| Previous Stroke/TIA | 14,818 (23.8%) | 314 (23.7%) | 14,504 (23.8%) | 0.933 |
| Carotid Stenosis | 1,112 (1.7%) | 21 (1.5%) | 1,091 (1.7%) | 0.577 |
| Atrial Fibrillation/Flutter | 10,203 (16.4%) | 208 (15.7%) | 9,995 (16.4%) | 0.495 |
| CAD/Prior MI | 9,815 (15.7%) | 195 (14.7%) | 9,620 (15.8%) | 0.291 |
| Chronic Renal Insufficiency | 6,489 (10.4%) | 167 (12.6%) | 6,322 (10.3%) | 0.008 |
| **Medications Prior to Admission** |  |  |  |  |
| Antithrombotic | 25,726 (42.1%) | 510 (39.0%) | 25,216 (42.1%) | 0.023 |
| Antiplatelets | 18,522 (29.5%) | 357 (26.9%) | 18,165 (29.5%) | 0.036 |
| Anticoagulants | 10,712 (17.%) | 234 (17.6%) | 10,478 (17.0%) | 0.574 |
| Antihypertensives | 29,480 (46.9%) | 647 (48.7%) | 28,833 (46.9%) | 0.182 |
| Cholesterol-Reducers | 23,484 (37.4%) | 476 (35.9%) | 23,008 (37.%) | 0.244 |
| Diabetic Medications | 10,826 (17.2%) | 298 (22.4%) | 10,528 (17.1%) | <.0001 |
| **Admission vital signs** |  |  |  |  |
| Heart Rate (30-200), bpm* | 84.9 (19.3) | 86.2 (19.3) | 84.9(19.3) | 0.021 |
| SBP (50-250), mm Hg* | 163.5 (34.6) | 159.3 (35.3) | 163.6 (34.6) | <.0001 |
| DBP (20-200), mm Hg* | 91.7 (23.3) | 89.3 (23.2) | 91.7 (23.3) | 0.001 |
| **Admission laboratory values** |  |  |  |  |
| Blood Glucose (20-800), mg/dL* | 151.60 (68.9) | 162.6 (78.2) | 151.3 (68.6) | <.0001 |
| HbA1c (0-20), %* | 6.2 (1.6) | 6.7 (2.0) | 6.2 (1.6) | <.0001 |
| LDL (30-500), mg/dL* | 94.8 (38.7) | 92.9 (36.3) | 94.9 (38.7) | 0.347 |
| Serum Creatinine (0-150), mg/dL* | 1.4 (4.0) | 1.5 (4.3) | 1.4 (4.0) | 0.065 |
| **Arrival Information** |  |  |  |  |
| Off-Hour Arrival (6PM to 7am) | 37,569 (59.8%) | 819 (61.7%) | 36,750 (59.8%) | 0.156 |
| Transferred in from other hospital | 23,421 (37.4%) | 526 (39.6%) | 22,895 (37.%) | 0.089 |
| Initial NIHSS Score (0-42)* | 11.9 (10.8) | 13.9 (10.7) | 11.9 (10.8) | <.0001 |
| Length of stay | 9.0 (11.7) | 12.8 (14.9) | 8.9 (11.6) | <.0001 |

We found evidence of significant differences in outcomes when comparing ICH patients with and without concomitant COVID-19 infection admitted during the pandemic (Table 2). In unadjusted regression analyses, ICH patients with versus without concomitant COVID-19 infection had 62% higher odds of poor outcome (OR 1.62, 95%CI 1.38-1.92), 48% higher odds of mortality (OR 1.48, 95%CI 1.31-1.67) and 41% higher odds of being discharged to a SNF/hospice (OR 1.41, 95%CI 1.23-1.61). Similar results were observed in multivariable regression analyses, where ICH patients with versus without concomitant COVID-19 infection had 68% higher odds of poor outcome (OR 1.68, 95%CI 1.41-2.01), 51% higher odds of mortality (OR 1.51, CI 1.33-1.71) and 66% higher odds of being discharged to a SNF (OR 1.66, 95%CI 1.43-1.93). Sensitivity analyses restricting the study population to 27,943 ICH cases enrolled at stroke centers (Supplementary Table 1) yielded similar results for all analyses, including poor outcome (OR 1.52, 95%CI 1.13-2.05), mortality (OR 1.19, 95%CI 0.97-1.47) and discharge to SNF (OR 1.66, 95%CI 1.33-2.01).

**Table 2.** The prevalence and odds ratios of outcomes are reported in COVID-19 positive patients compared to those with no documented infection (reference).

| **Stroke Outcome at Discharge** | **Model Type** | **N Available data** | **Odds Ratio** | **Confidence Interval** |
| --- | --- | --- | --- | --- |
| **Discharge to SNF or hospice** | Adjusted<sup>B</sup> | 47,693 | 1.66 | 1.43 - 1.92 |
|  | Unadjusted | 49,167 | 1.40 | 1.23 - 1.60 |
| **Length of stay<sup>A</sup>** | Adjusted<sup>B</sup> | 58,481 | 1.32 | 1.25 - 1.39 |
|  | Unadjusted | 60,278 | 1.43 | 1.35 - 1.51 |
| **High mRS (4, 5, 6)** | Adjusted<sup>B</sup> | 39,668 | 1.68 | 1.40 - 2.01 |
|  | Unadjusted | 40,718 | 1.62 | 1.37 - 1.91 |
| **Mortality** | Adjusted<sup>B</sup> | 60,894 | 1.50 | 1.33 - 1.71 |
|  | Unadjusted | 62,743 | 1.47 | 1.31 - 1.66 |

<sup>A</sup> The estimate is a relative risk (with its 95% CI); not odds ratio when the outcome is length of stay.
<sup>B</sup> The following covariates were used in the adjusted models: age, sex, race/ethnicity, antiplatelet or anticoagulant prior to admission, Hypertension, Diabetes, Heart failure, CAD/MI, Prior stroke, Renal insufficiency, Atrial Fibrillation, Smoking, Alcohol/Drug Abuse, Sleep apnea, Pre-stroke mRS, Anticoagulant, Antiplatelet, Hypertensive, arrival via EMS, arrival on vs. off hours, admission NIHSS score, Antiplatelet, Anticoagulant, region, teaching hospital, number of beds, annual stroke volume, rural location, stroke center status

### Differences between ICH patients admitted before and during the pandemic

The second stage of the present study, focused on comparing ICH cases that took place before and during the COVID-19 pandemic, evaluated a total of 127,424 ICH patients (mean age 67.7 [SD 15.3], female sex n=59,270 [46.5%]), including 62,743 during the pre-pandemic period and 64,681 during the pandemic (Table 3). The baseline characteristics of ICH patients admitted before and during the pandemic were overall similar, despite some statistically significant results expected given the large sample size but that corresponded to small differences (Table 3).

**Table 3.**
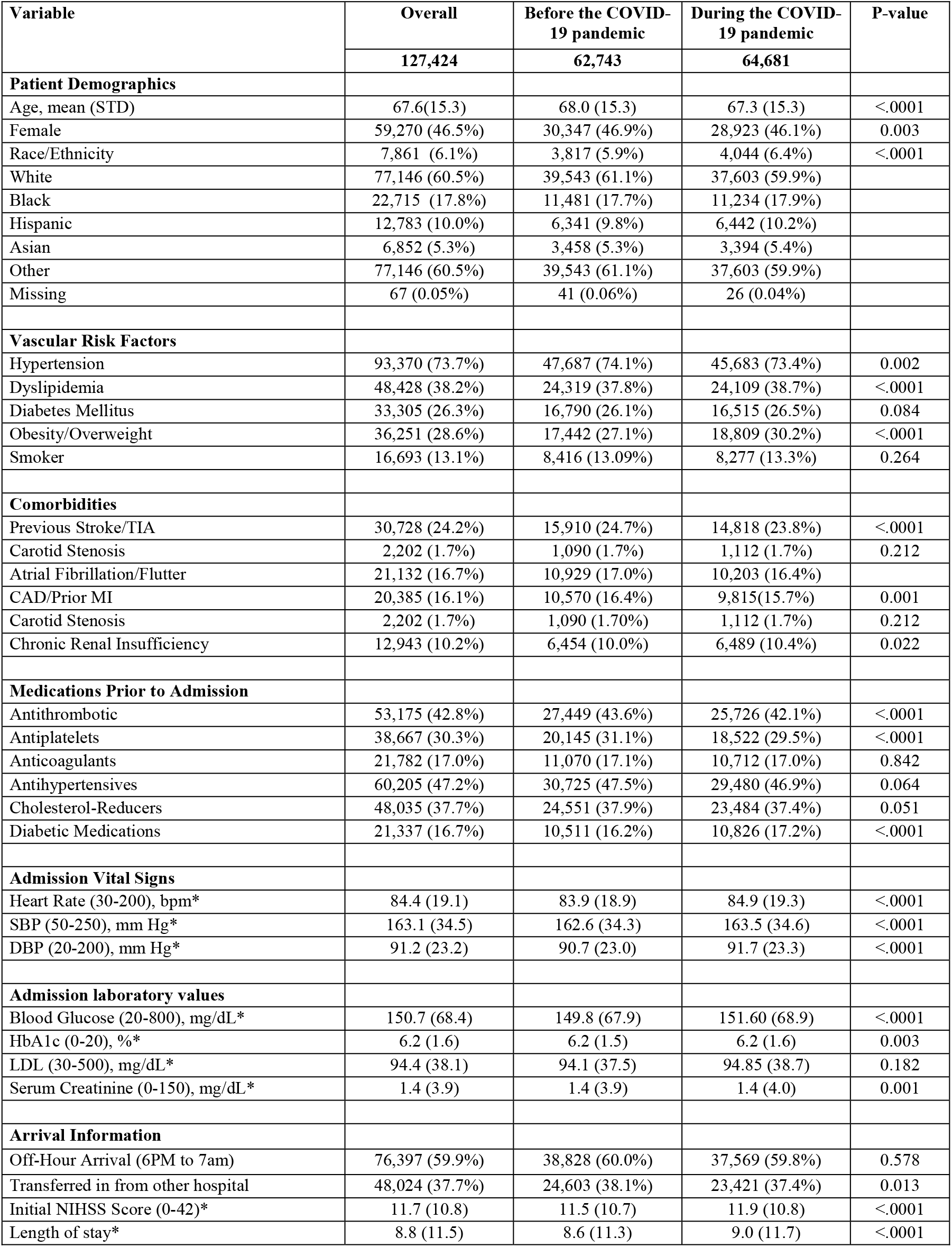
Description and comparison of patient baseline characteristics data among ICH patients who were admitted to a hospital before the COVID-19 pandemic and time-matched patients during the COVID-19 pandemic.

We also found evidence of significant differences in outcomes when comparing ICH patients admitted before and during the pandemic (Table 4). In unadjusted regression analyses, ICH patients admitted during versus before the COVID-19 pandemic had a 5% higher odds of poor outcome (OR 1.05, 95%CI 1.01-1.08) and less odd of being discharge to an to SNF/hospice (OR 0.90, 95%CI 0.88-0.93). No statistically significant differences observed in mortality (OR 1.00, 95%CI 0.98-1.03) or discharge Similar results were observed in multivariable regression analyses that indicated that ICH patients admitted during versus before the COVID-19 pandemic had a 10% higher odds of poor outcome (OR 1.10, 95%CI 1.07-1.14), with no significant changes observed in discharge to SNF/hospice (OR 0.93, 95%CI 0.90-0.95). In contrast to univariate analyses, multivariable regression demonstrated higher odds of mortality (OR 1.05, 95%CI 1.02-1.08). Sensitivity analyses restricting the study population to 57,787 ICH cases enrolled at comprehensive stroke centers (Supplementary Table 2) yielded comparable results, including a significant association for poor outcome (OR 1.14, 95%CI 1.08-1.21) and no statistically significant differences for mortality (OR 1.03, 95%CI 0.98-1.07).

**Table 4.**
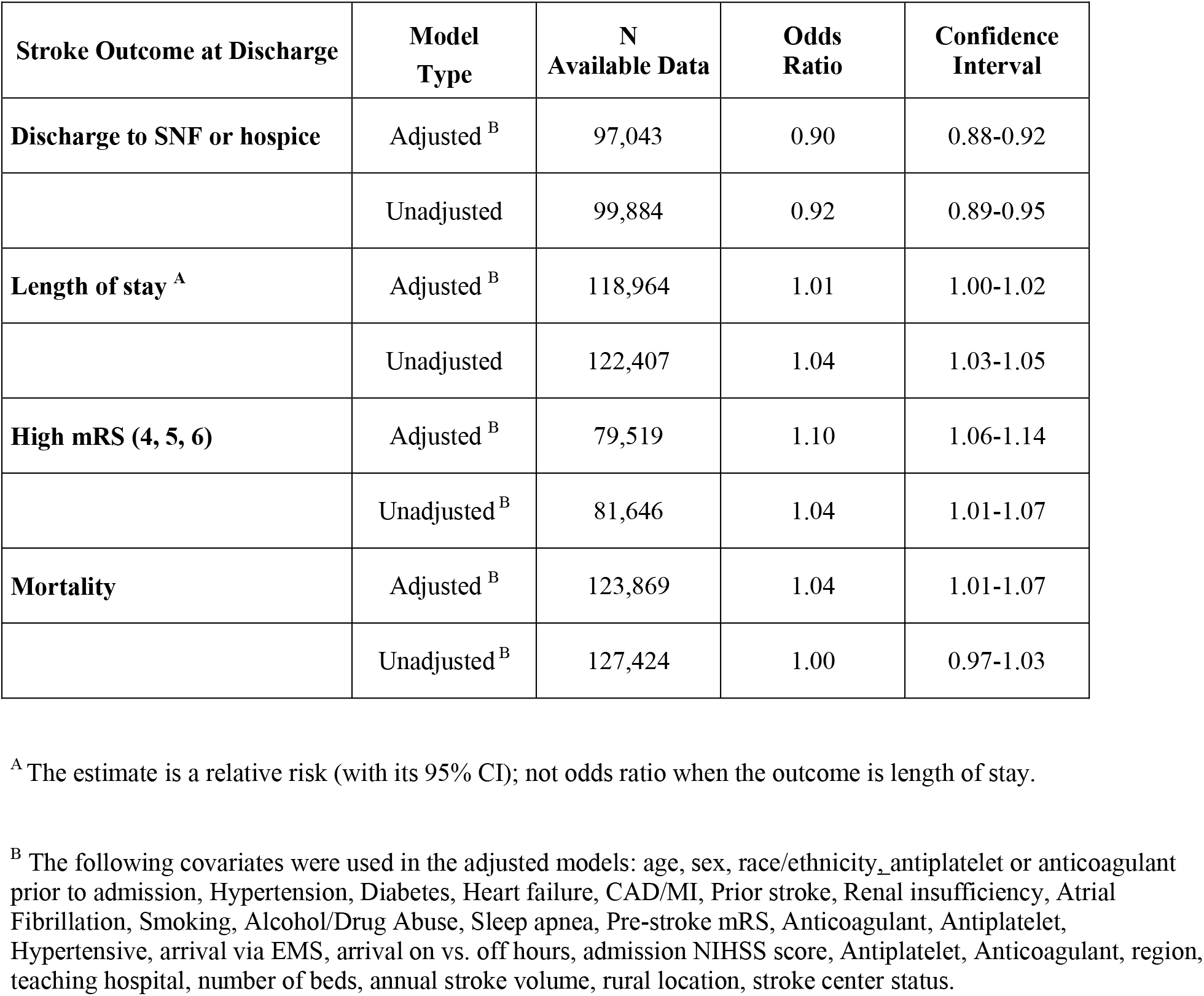
The prevalence and odds ratios of outcomes are reported during the pandemic vs. before (reference) the COVID-19 pandemic.

## DISCUSSION

We present the results of a large observational study aimed at describing the consequences of the COVID-19 pandemic in the outcomes of patients with spontaneous, non-traumatic ICH. First, we compared the outcomes of ICH patients with and without concomitant COVID-19 infection admitted during the pandemic, finding that those with concomitant infection had higher risk of poor functional outcome, death and discharge to SNF or hospice. Second, we compared ICH patients admitted during a similar time period before and during the pandemic, finding that the latter had a slightly higher risk of poor functional outcome and death. Sensitivity analyses restricting the analyses to ICH patients admitted to stroke centers yielded comparable results. We subsequently compared the outcomes of ICH patients admitted before and during the pandemic. We found that, during the pandemic, ICH patients had a higher risk of poor outcome when compared to those admitted before the pandemic. Of note, the size of this association (10% higher risk of poor outcome) was smaller than that observed when comparing ICH patients with and without COVID-19 infection admitted during the pandemic (around 50%).

Mounting evidence indicates that the COVID-19 infection negatively impacts the outcomes of several cardiovascular diseases. As an example, a retrospective study based on the National Inpatient Sample that evaluated outcomes in ST-segment elevation myocardial infarction, patients with COVID-19 infection (n=5,786) had 47% higher mortality than those who were not infected (n=166,399).^15^ A similar study focused on acute ischemic, that used the Vizient Clinical Data Base to evaluate 2,086 COVID-19+ and 166,586 COVID-19 stroke patients, found significant higher unadjusted mortality rates in the later (30.4% versus 6.5%, respectively).^16^ Despite this compelling evidence for a role of the COVID-19 infection as modifier of the natural history of acute cardiovascular syndromes, the existing data on hemorrhagic stroke in general, and ICH in particular, remains limited to small, single-center studies with limited statistical power to effectively identify differences introduced by the COVID-19 infection.^17,18^ One previous study showed that patients with spontaneous ICH or SAH and comorbid COVID infection were more likely have diabetes been obese and to have higher rates of death when compared with controls.^8^

Our study provides important novel findings that directly tackle this knowledge gap. The first stage of our study, focused on ICH patients admitted during the pandemic and aimed at comparing those with and without concomitant COVID-19 infection, identified several differences between the groups. We found that ICH patients who were COVID-19-positive were younger and more likely to belong to underrepresented race/ethnic groups (Black or Hispanic). These findings are in line with prior studies that found similar results when evaluating patients who had sustained an ischemic stroke.^17,8,4^ In addition, we found that ICH patients who were COVID-19-positive had worse clinical trajectories, as evidenced by higher risk of poor functional outcome, higher mortality and higher risk of being discharged to SNF/hospice. The design and available data of this study does not allow us to identify the reasons for this worse clinical trajectory. However, based on published of COVID-19 infection in other diseases, it is reasonable to speculate that the concomitant contribution of severe respiratory disease and/or acute thromboembolic episodes triggered by accelerated inflammatory responses, both triggered by the virus, could be responsible for the observed worse outcomes.^19,20,21^ Given that inflammation plays an important role in several biological mechanisms linked to secondary brain injury that takes place around the brain hemorrhage^22^, it is also possible that heightened responses triggered by the virus could mediate these poor outcomes.

The results of the second stage of this study, comparing ICH patients admitted before and during the pandemic, indicate that factors other than the biological changes introduced by the infection are likely to also play a role in causing the observed differences. We found that ICH patients admitted during the pandemic had higher risk of poor functional outcome and death, although the size of these associations were smaller than those observed when comparing COVID-19 positive versus negative admitted during the pandemic. These findings emphasize the need to understand and tackle aspects that go beyond the possible biological effects of the infection, including access to care and changes in standards of care during the pandemic. An example of these changes in patterns of care is our finding ICH patients during COVID-19 pandemic were more frequently admitted to hospitals with a lower volume of ischemic strokes and ICH stroke admission and lower numbers of beds.

The main strengths of our study are the uniquely large sample size, allowing the appropriate evaluation of clinical and biological differences in these patients; the multi-institutional design, that increases the generalizability of our findings; and the pre-specified data collection strategy, that allows the effective harmonization of data across the multiple institutions involved. A number of limitations should also be mentioned. First, the universe of hospitals participating in GWTG-Stroke may not be representative of the entire universe of hospitals in the US limiting the applicability of these findings to some settings, like rural areas and small towns. Second, we lacked detailed data on the timing of COVID-19 diagnosis during the hospital admission, the specific diagnostic study used diagnose the infection, and the treatments that were implemented to tackle the infection. Third, we did not have granular data on the specific clinical complications that occurred during the admission and could shed light on the specific clinical reasons for the observed worse outcomes. There is also the likelihood of residual, unmeasured confounding.

In summary, our findings indicate that, during the pandemic, ICH patients with concomitant COVID-19 infection had worse functional outcomes and mortality when compared to those without the infection. Similar findings, but with smaller effects sizes, were observed when comparing ICH patients admitted before and during the pandemic. These results extend the findings of similar studies focused on other acute cardiovascular pathologies, including ischemic stroke, to spontaneous, non-traumatic ICH. These novel findings support further research focused on understanding the biological and clinical mediators that lead to the observed worse outcomes and identifying risk-prediction strategies to identify ICH patients at high risk of poor outcomes when facing a concomitant COVID-19 infection.

## Data Availability

Get With The Guidelines is a proven in-hospital approach to improving patient outcomes across cardiovascular and stroke focus areas. Access to the data is made through the Precision Medicine Platform. Previous approval of a proposal is needed to be granted access.

https://www.heart.org/en/professional/quality-improvement/get-with-the-guidelines/get-with-the-guidelines-stroke

## ACKNOWLEDGEMENTS

The Get With The Guidelines programs are provided by the American Heart Association. The American Heart Association Precision Medicine Platform (https://precision.heart.org/) was used for data analysis. IQVIA (Parsippany, New Jersey) serves as the data collection and coordination center.

